# Impact of prenatal exome sequencing for fetal genetic diagnosis on maternal psychological outcomes and decisional conflict in a prospective cohort

**DOI:** 10.1101/2020.08.05.20169029

**Authors:** Asha N. Talati, Kelly L. Gilmore, Emily Hardisty, Anne D. Lyerly, Christine Rini, Neeta L. Vora

**Author notes:** Corresponding Author: Asha N. Talati MD MS, 3010 Old Clinic Building, Campus Box 7516, Chapel Hill, NC, 27599.

## Abstract

**Objective:** To evaluate associations between prenatal trio exome sequencing (trio-ES) and psychological outcomes among women with an anomalous pregnancy.

**Methods:** Trio-ES study enrolling patients with major fetal anomaly and normal microarray. Women completed self-reported measures and free response interviews at two time points, pre- (1) and post- (2) sequencing. Pre-sequencing responses were compared to post-sequencing responses; post-sequencing responses were stratified by women who received trio-ES results that may explain fetal findings, secondary findings (medically actionable or carrier couple status), or negative results. Free responses were content analyzed.

**Results:** 115 trios were enrolled. Of those, 41/115 (35.7%) received results from trio-ES, including 36 (31.3%) who received results that may explain the fetal phenotype. These women had greater post-sequencing distress compared to women who received negative results, including generalized distress (p=0.03) and test-related distress (p=0.2); they also had worse psychological adaptation to results (p=0.001). Genomic knowledge did not change from pre- to post-sequencing (p=0.51). Major themes from content analyses included *closure, future pregnancy, altruism, anxiety*, and *gratitude*.

**Conclusions:** Women show more distress after receiving trio-ES results compared to those who do not.

## Introduction

Fetal anomalies affect 3-5% of all pregnancies and account for nearly 20% of perinatal mortality.^1^ Advances in prenatal genetic screening and diagnosis, ranging from chromosomal microarray to next-generation sequencing strategies, offer an opportunity to provide information to inform reproductive decision making and more tailored pre- and post-natal pregnancy management. Further, these advances allow for multi-disciplinary care planning, and they inform and improve neonatal care and health outcomes.^2^ Prenatal trio exome sequencing (trio-ES) is one such advance. Evidence thus far suggests that trio-ES increases diagnostic yield when standard genetic testing (karyotype and microarray) is normal.^3 4 5 6^ The two largest studies found a diagnostic rate of 8.5% and 10%, overall, and 15.4% and 19% in cases of multisystem abnormalities.^4 5^ Given emerging evidence showing improve diagnostic rates and the generally rapid uptake of next generation sequencing strategies in clinical care, it is likely that trio-ES will eventually be integrated into prenatal clinical care.

However, to date, there are no large studies with prospectively collected longitudinal data assessing women’s understanding of trio-ES results, the psychological impact of testing, and influence on future reproductive decisions. Rapid integration of new technologies such as exome sequencing pose significant and unique ethical and counseling challenges.^7^ Women’s ability to understand and accept information from next generation sequencing technologies such as trio-ES is likely to depend on factors that influence these responses to other diagnostic strategies. For instance, these factors may include health literacy, socioeconomic status, race/ethnicity, cultural and religious beliefs, attitudes towards termination, experiences with disability, and the methods and mode of information delivery.^8 9 10 11 12 13^Furthermore, genetic information provided in this setting may be prone to uncertainty (e.g., variants of uncertain significance) or they may have strong potential to impact parental well-being (e.g., medically actionable secondary findings). Thus, psychosocial adjustment and the behavioral impact of prenatal exome sequencing results are important outcomes to understand prior to integration of trio-ES into the diagnostic algorithm. Because of the limited empiric data available to guide best counseling and implementation of trio-ES in this arena, our objective was to understand the association between sequencing outcomes and maternal decisional conflict, psychological adaptation, and future reproductive decisions by leveraging our existing cohort from the UNC-Chapel Hill fetal exome sequencing project. We also evaluated genomics knowledge pre- and post- counseling to assess whether baseline genetic knowledge or knowledge gain during counseling may contribute to differences in psychological outcomes. We hypothesized the following: (1) women who received a result from trio-ES that may explain the fetal phenotype would have higher levels of generalized distress and worse psychological adaptation to their results than women who received secondary findings or no reportable results; (2) higher maternal educational level and baseline genomic knowledge would be associated with lower generalized distress and better psychological adaptation post-sequencing, and (3) pre-sequencing genomic knowledge would be higher among women who self-identify as having higher levels of education, income, and prior experience with genetic screening or testing.

## Materials and Methods

The University of North Carolina at Chapel Hill institutional review board (13-4084) provided approval for this study. We enrolled trios (parents and fetus) in pregnancies complicated by either isolated or multiple congenital anomalies with a normal karyotype and microarray. Patients were identified from prenatal diagnosis clinics from the University of North Carolina (UNC) at Chapel Hill (Chapel Hill, NC) and from prenatal diagnosis clinics across the United States between July 2014 and December 2019. A certified prenatal genetic counselor performed consent procedures for the study, and non-local families gave consent via secure web-based video calls. Mothers and fathers provided consent separately so that counselors could discuss the chance of non-paternity with mothers and allow them to opt out of the study if they desired. Trios were included in the study if the following inclusion criteria were met: 1) singleton gestation; 2) suspected genetic etiology of congenital anomalies visualized on ultrasound; 3) lack of diagnosis after karyotype, microarray, and if indicated, gene-specific sequencing; and 4) presence of DNA from fetus, mother, and father. Parent-fetus trios were identified both prospectively and retrospectively and were enrolled at various gestational ages or after the pregnancy was completed. Trios enrolled prospectively were only approached for recruitment to the study after they had received standard clinical counseling and had made a decision about pregnancy management (e.g., continuation versus termination of pregnancy). The study was not mentioned prior to this time to avoid impacting parental decision-making. Trios were also identified retrospectively through the UNC Perinatal Database, a repository of patients who received prenatal and delivery care at UNC (1996 to present). Women who previously indicated a desire to be re-contacted for additional fetal testing and who had fetal cells archived and available for DNA extraction were also approached for enrollment.

After enrollment, participants had pretest counseling by a certified prenatal genetic counselor regarding trio-ES and the possible results it can provide. All participants agreed to learn findings that explained the fetal phenotype, medically actionable secondary findings in a parent, and/or carrier couple status for significant autosomal recessive conditions. Participants who chose to opt out of learning any of the former findings at the time of consent were excluded from the study. After enrollment, we obtained maternal and paternal blood and extracted DNA in the Biospecimen Processing Facility, a core UNC-CH laboratory. Fetal DNA was extracted from stored specimens, such as amniocytes, chorionic villi, or umbilical cord blood as appropriate. For non-local cases, we directly received extracted fetal DNA from the outside institution. All genetic variants that were identified were confirmed by Sanger sequencing at the UNC Molecular Genetics Laboratory (MGL), a CLIA-certified and CAP-accredited facility, using a duplicated sample. After confirmation of results, parents were given the option to sign a separate consent form to have their own or their child’s variant placed in the medical record.

Data were collected at two time points. Immediately after pre-sequencing counseling and enrollment, the mother completed a pre-sequencing demographics questionnaire and a measure of genomic knowledge adapted from the NCGENES study (UNC-GKS). This measure includes 25 statements that participants are asked to judge as being true or false. Possible responses included “don’t know/unsure” to reduce guessing. The statements were designed to evaluate recall and understanding of new information received in the study. Correct responses were summed to create a score ranging from 0 (no answers correct) to 25 (all answers correct) [Supplemental material 1].^14^ Women also completed a validated measure of generalized distress (symptoms of anxiety and depression), the Hospital Anxiety and Depression Scale (HADS)^15^, and were asked an open-ended question prompting them to describe their expectations of and reasons for pursuing exome sequencing for fetal diagnosis (Figure 1). After return of results, participants were counseled regarding findings from trio-ES and repeated the measures of genomic knowledge and generalized distress (post-sequencing measures). They also completed the following assessments: (1) test-related distress (adapted from the Multidimensional Impact of Cancer Risk Assessment (MICRA);^16^ (2) decisional conflict using the Decisional Conflict Scale (DCS);^17^ (3) psychological adaptation to results (perceptions of non-intrusiveness, support, self-worth, certainty, and self-efficacy) using the Psychological Adaptation to Genetic Information Scale (PAGIS);^18^ and (4) brief, open ended question prompting women to summarizing their interpretation of findings from trio-ES and experience with sequencing in their own words. The post-sequencing assessment was completed 2 weeks after sequencing results were delivered. Interviews and assessments were completed in person, on-line, by phone, or by web-based video platform dependent on the participant location (Figure 1).

**Figure 1.**
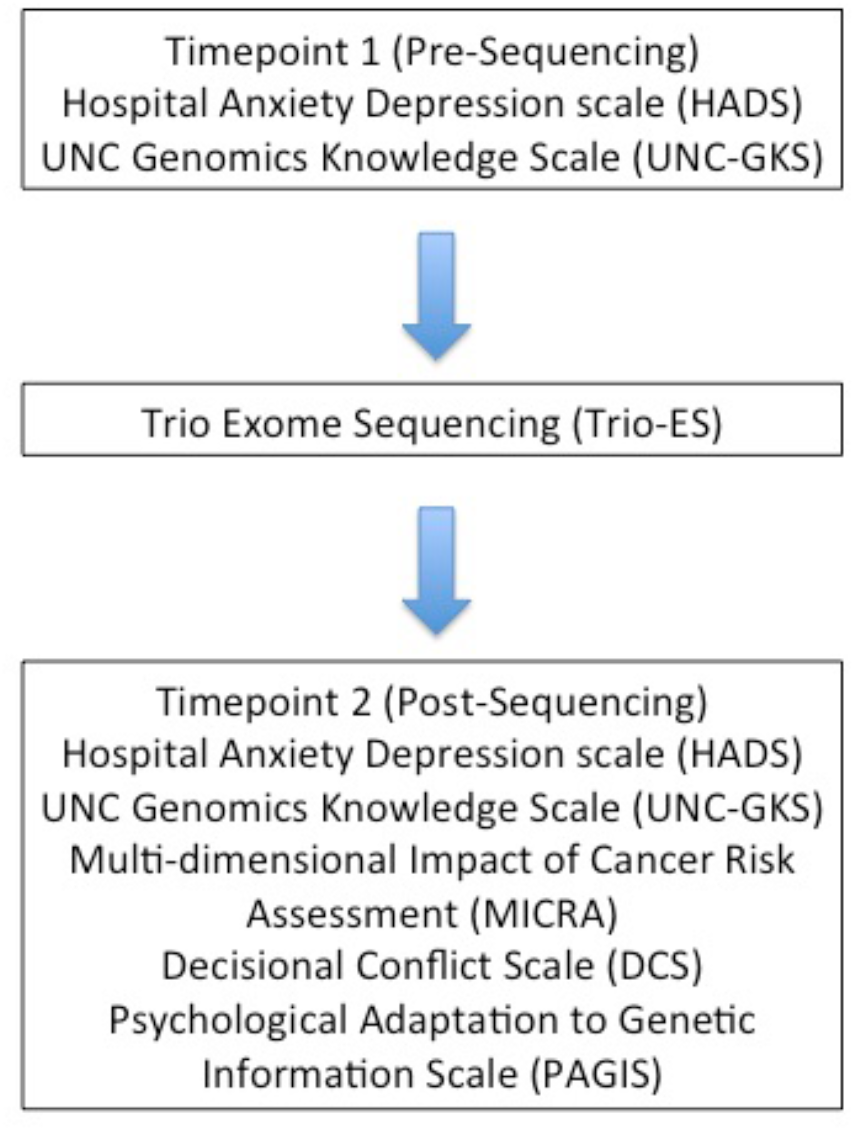
Study design and surveys included at time-points 1 and 2 (pre and post sequencing)

Participants’ demographic characteristics were analyzed using descriptive statistics. Pre- and post-sequencing genomic knowledge and generalized distress were compared. Post-sequencing generalized distress, test-related distress, decisional conflict, and psychological adaptation to results were compared for the following groups: (1) women who received negative results from trio-ES, (2) women who received results that may explain the fetal phenotype; and (3) women who received a report of medically actionable secondary findings or carrier couple results. Additionally, associations between pre-sequencing genomic knowledge test scores, maternal demographic characteristics, and post-sequencing self-reported outcomes were assessed. Bivariable analyses were completed using t-test, Wilcoxon rank-sum, Kruskal Wallis test, Spearman’s correlation, and linear regression, as appropriate. Pairwise comparisons using Tukey’s method was employed as appropriate. P <0.05 was considered statistically significant.

All analyses were completed using STATA 15 statistical software (College Station, TX). Open-ended responses at Times 1 and 2 were analyzed using a thematic analysis approach with an inductive coding style.^19^ Common themes were identified and developed into a code book. Two researchers were involved in transcription and coding to ensure inter-coder reliability. Open-ended responses were reviewed in an iterative fashion, and after primary and secondary coding consensus was reached regarding major emerging themes and subthemes. Dedoose Version 8.0 (Los Angeles, CA) was used for coding and analysis.

## Results

One hundred fifteen trios were enrolled in the study. Of these participants, 41 (35.7%) received results from trio-ES, of which 36 (31.3%) received results that may explain the fetal phenotype. Seven (6.1%) received secondary findings (either medically actionable incidental findings in a parent or carrier status for significant autosomal recessive conditions). Maternal demographic characteristics are shown in Table 1. Median age of women enrolled in the study was 31 (IQR 27,34). Most of the cohort self-identified as Non-Hispanic White (71.3%), married (84.3%), and as having some college education (87%). In addition, over half the cohort has had prior experience with prenatal genetic screening or diagnosis (57.4%). Fetal genetic material for diagnosis was obtained through CVS, amniocentesis, cord blood, or products of conception. The majority of fetal genetic material was obtained from amniocentesis or CVS (70.4%).

**Table 1.** Demographic characteristics of women in cohort.

|  |  |
| --- | --- |
|  | N=115 |
| Age (median IQR) | 31 (27,34) |
| Self-Identified Race |  |
| Non-Hispanic White | 82 (71.3) |
| Black | 8 (6.9) |
| Hispanic | 15 (13.0) |
| Asian or Pacific Islander | 7 (6.0) |
| Native American/Alaskan | 2 (1.7) |
| Other | 8 (7.0) |
| Married | 97 (84.3) |
| Education |  |
| Less than high school | 2 (1.7) |
| High school graduate | 10 (8.7) |
| Partial college | 14 (12.2) |
| 2 year college | 16 (13.9) |
| 4 year college | 54 (47.0) |
| Graduate degree | 16 (13.9) |
| Employed | 72 (62.6) |
| Income > \$90,000 | 58 (50.4) |
| Prior Genetic Testing | 66 (57.4) |

\*Data are presented as n(%) unless noted otherwise

Of the 115 women who participated in trio-ES, 101 (88%) completed the post-sequencing assessment. Overall, women who received results that may explain the fetal phenotype had higher post-sequencing generalized distress, test-related distress, and worse psychological adaptation to testing compared to women who received medically actionable secondary findings or negative trio-ES results. Higher scores on each of these scales are suggestive of the following: increased symptoms of anxiety and depression (HADS), test-related distress and uncertainty (MICRA), and intrusive thoughts and lack of support (PAGIS). Specifically, women who received findings that may explain the fetal phenotype or who received medically actionable secondary findings reported higher test-related distress (MICRA) than women who received negative results (8.1 vs. 4.8 vs. 3.1, p<0.001). In pairwise comparisons, test related distress was significantly higher amongst the group of women who received findings that may explain the fetal phenotype compared to those receiving negative results (mean difference = 5.02, p<0.001). There was no difference in likelihood to report positive experience with testing on the MICRA between groups. Similar patterns appeared for several subscales of the measure of psychological adaptation to results (PAGIS), including intrusive thoughts (8.3 (SD 5.8) vs. 8.7 (SD 6.7) vs. 4.5 (SD 5.9), p=0.003) and difficulty with support from family and peers (9.3 (SD 9.2) vs. 7 (SD 4.9) vs. 3.7 (SD5.0), p<0.001). Amongst pairwise comparisons, intrusive thoughts, difficulty with support, and overall worse adaptation to results were significantly different among women who received results that explained the fetal phenotype than those who received negative results (mean difference = 3.7, p=0.01; mean difference = 5.5, p<0.001, mean difference = 8.7 p=0.005, respectively) (Table 2).

Women who received results that explain the fetal phenotype had significantly higher overall score on the HADS (10.1 (SD 6.5), 6.5 (SD 5.7), 7.2 (SD 5.2) p=0.03), specifically the depression subscale (3.2 (SD 3.3), 1.2 (SD 2.5), 12. (SD 2.7), p=0.003), when compared to women who received medically actionable secondary findings or negative results. Pairwise comparisons demonstrated that there was a significant difference in the depression subscale between women who received results that explain the fetal findings compared to women that received negative results (mean difference = 1.97, p = 0.003) Notably, women who received results did not differ in their decisional conflict when compared to women who had negative results, indicating that they did not differ in their perceptions of feeling informed, clear about their values, and satisfied with their choice (DCS scores 41.8 (SD 5.5) vs. 43.2 (SD 5.4) vs. 40 (SD 0) p=0.34). Similarly, there was no difference in mean DCS scores between groups when employing pairwise comparisons (p= 0.28, 0.47, 0.23). (Table 2).

**Table 2.** Post-sequencing survey results stratified by receipt of fetal trio WES results, receipt of medically actionable secondary finding, or receipt of no results.

|  | **No trio-WES**<br>**results reported** | **Fetal trio-WES results**<br>**reported** | **Secondary**<br>**findings reported** | **P-value** |
| --- | --- | --- | --- | --- |
|  | **N=61** | **N=27** | N=5 |  |
| **HADS** |  |  |  |  |
| **Anxiety Subscale** | 5.3 (3.7) | 6.9 (4.0) | 6 (3.7) | 0.17 |
| **Depression Subscale** | 1.2 (2.5) | 3.2 (3.3) | 1.2 (2.7) | 0.003 |
| **Total Score** | 6.5 (5.7) | 10.1 (6.5) | 7.2 (5.2) | 0.03 |
|  | **N=67** | **N=28** | N=6 |  |
| **MICRA** |  |  |  |  |
| **Distress** | 3.1 (5.2) | 8.1 (6.2) | 4.8 (4.9) | 0.0003 |
| **Uncertainty** | 5.2 (6.2) | 6.1 (4.7) | 5.7 (4.6) | 0.39 |
| **Positive Experience** | 10.1 (4.3) | 9.6 (3.4) | 7.8 (4.5) | 0.28 |
| **Total** | 29.3 (16.5) | 36.3 (13.3) | 27.2 (18.1) | 0.02 |
|  | **N=61** | **N=27** | N=5 |  |
| **Decision Conflict Scale** | 41.8 (5.5) | 43.2 (5.4) | 40 (0) | 0.34 |
|  | **N=61** | **N=24** | N=6 |  |
| **PAGIS** |  |  |  |  |
| **Non-obtrusiveness** | 4.5 (5.9) | 8.3 (5.8) | 8.7 (6.7) | 0.003 |
| **Support** | 3.7 (5.0) | 9.3 (9.2) | 7 (4.9) | 0.0007 |
| **Certainty** | 2.5 (4.0) | 1.9 (2.8) | 1.3 (1.6) | 0.92 |
| **Total** | 10.7 (12.3) | 19.4 (13.9) | 17 (10.2) | 0.001 |

\*Data are presented as mean (SD) unless noted otherwise
\* HADS is based on a scale of 0-21 with score > 8 considered a positive screen. Anxiety and Depression domains can be scored individually with scores >7 considered a positive screen (Stern Occupational Med 2014). MICRA is a 25-question scale scored (0-75) with subscales listed here. Decisional Conflict Scale (5-item) scored 13-17, >15 concerning for decisional conflict. PAGIS was adapted to an 18-item scale with subscales shown here.

Mean generalized distress scores were significantly higher among women at pre-sequencing than post-sequencing, both in terms of the overall score and when we conducted follow up analyses evaluating the anxiety and depressive symptoms subscales of this measure separately (HADS pre-sequencing mean anxiety scores 7.8 (SD 4.2), post sequencing mean anxiety scores 5.8 (SD 3.8), p=0.005; HADS pre-sequencing mean depression scores 4.2 (SD 4.1), post sequencing mean depression scores 1.8 (SD 2.9), p<0.001; HADS pre-sequencing mean total score 11.9 (SD 7.3), post-sequencing mean total score 7.6 (SD 6.1), p<0.001). There was no significant correlation between post-sequencing psychological outcomes and maternal demographic characteristics such as age, race, marital status, income, educational level, and prior genetic testing. However, receipt of any trio-ES result (either one that may explain the fetal phenotype or a secondary finding) was associated with higher post-sequencing test-related distress (7.2 SD (1.0) vs. 3.2 (SD 0.66), p<0.001), higher generalized distress (9.4 (SD 1.1) vs. 6.5 (SD 0.75), p=0.02) and worse psychological adaptation (18.2 (SD 2.3) vs.10.8 (SD 1.6), p=0.0007) scores.

Within the cohort overall, mean genomic knowledge scores did not change from pre- to post-sequencing (pre-sequencing mean number correct 21.7 (out of 25) (SD 0.22), post-sequencing mean number correct 21.9 (SD 0.32), p=0.51). Pre-sequencing genomic knowledge was associated with higher education level (p=0.0013). Mean genomic knowledge score among women who did not complete high school was 13.5 correct (out of 25) (SD 1.5) whereas women with graduate degrees had a mean score of 22.8 correct (SD 0.6). Regression analysis demonstrated that women with higher education levels (adjusted coefficient 0.37, p<0.001) and previous genetic testing (adjusted coefficient 0.02, p<0.001) were more likely to have higher genomic knowledge scores at the pre-sequencing assessment (model adjusted for age, race, marital status, employment, income level). Of note, there was no significant association between pre-sequencing genomic knowledge and post-sequencing psychological outcomes.

The majority of women in our cohort pursued trio-ES in hopes of having future children (83%), information to guide prenatal diagnosis for future pregnancies (90%), and reassurance to reduce concerns about future children (90%). A significant proportion of women sought trio-ES for a definite explanation for findings in the current pregnancy (90%), despite pre-test counseling regarding diagnostic yield. Post-sequencing, 70% of women in the cohort reported that trio-ES outcomes did not change their future reproductive plans and nearly half the cohort remained undecided about future pregnancy (47.9%) (Table 3).

**Table 3.**
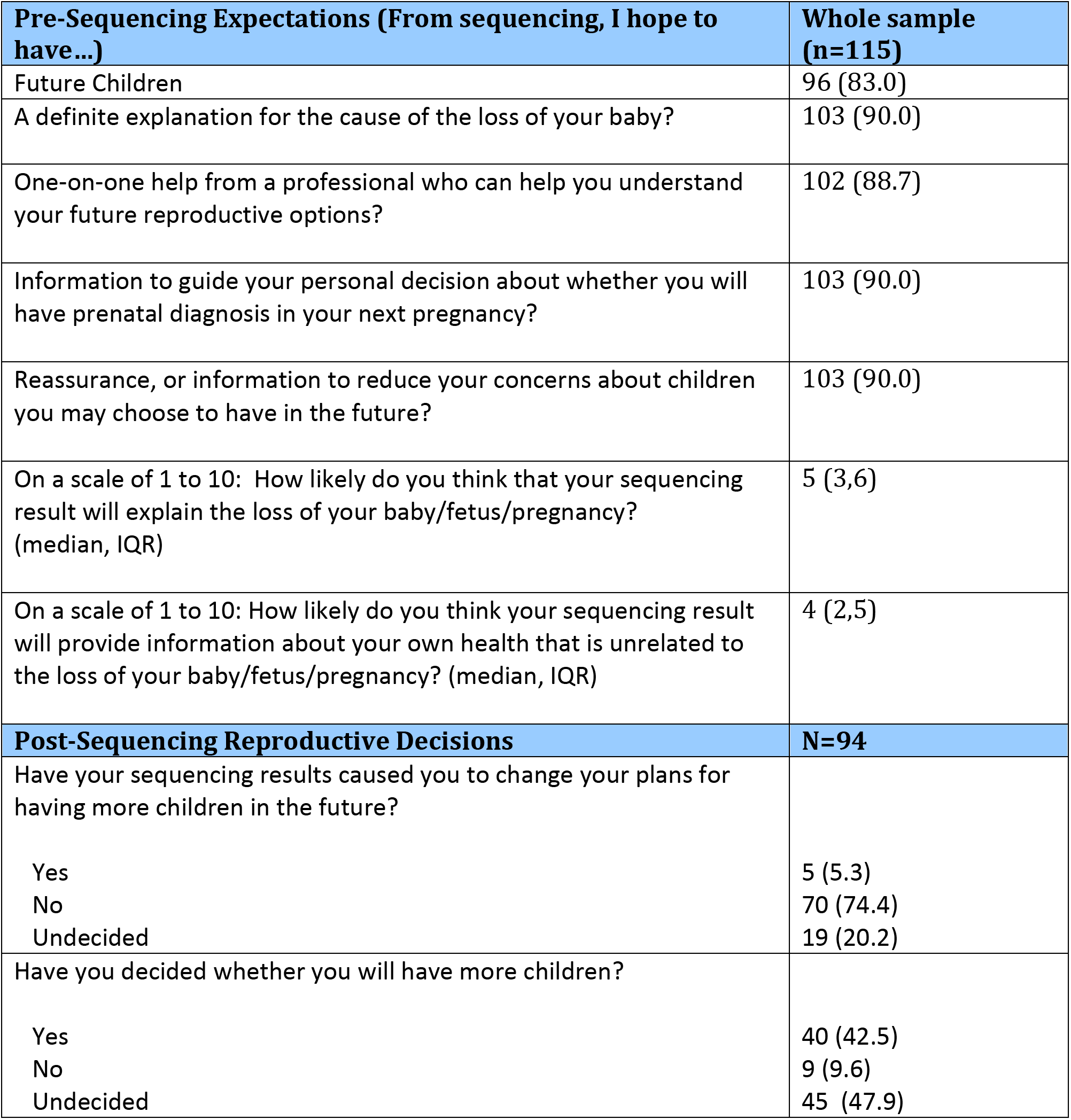
Expectations of WES and pre sequencing and reproductive decisions postsequencing (Data are shown as n(%) unless noted otherwise.)

| **Pre-Sequencing Expectations (From sequencing, I hope to have...)** | **Whole sample (n=115)** |
| --- | --- |
| Future Children | 96 (83.0) |
| A definite explanation for the cause of the loss of your baby? | 103 (90.0) |
| One-on-one help from a professional who can help you understand your future reproductive options? | 102 (88.7) |
| Information to guide your personal decision about whether you will have prenatal diagnosis in your next pregnancy? | 103 (90.0) |
| Reassurance, or information to reduce your concerns about children you may choose to have in the future? | 103 (90.0) |
| On a scale of 1 to 10: How likely do you think that your sequencing result will explain the loss of your baby/fetus/pregnancy? (median, IQR) | 5 (3,6) |
| On a scale of 1 to 10: How likely do you think your sequencing result will provide information about your own health that is unrelated to the loss of your baby/fetus/pregnancy? (median, IQR) | 4 (2,5) |
| **Post-Sequencing Reproductive Decisions** | **N=94** |
| Have your sequencing results caused you to change your plans for having more children in the future? |  |
| Yes | 5 (5.3) |
| No | 70 (74.4) |
| Undecided | 19 (20.2) |
| Have you decided whether you will have more children? |  |
| Yes | 40 (42.5) |
| No | 9 (9.6) |
| Undecided | 45 (47.9) |

Pre-sequencing open-ended responses revealed three major themes informing maternal decision to pursue trio-ES in the context of our study: *altruism, closure*, and *future planning*. Many women who noted pursuing sequencing “to help other families” or to find “an answer” prefaced their reasoning with having few personal expectations from trio-ES (46%). When discussing interpretation of sequencing results, major emergent themes included *uncertainty* and *gratitude*. Among the 27 women who expressed feelings of uncertainty after trio-ES, the majority received negative results from sequencing (85.2%). Fifteen women expressed feelings of gratitude post sequencing, mentioning “closure” and “peace of mind.” Among those women, 60% received some result from trio-ES (9/15). The remaining 6 women received negative results; however, they cited feelings of relief associated with having received negative findings. Table 4 provides example quotes for emergent themes pre and post sequencing in our cohort.

**Table 4.** Representative Quotations by Theme.

| Theme | Quotation |
| --- | --- |
| Pre-Sequencing |  |
| Altruism | 1. *I don't expect to get any kind of result. I just want to help future families. Have something to "name" this with.*<br>2. *I don't really expect to get an answer. We are more doing it for research purposes to help other families.* |
| Closure | 1. *I hope to understand her issues with genetics but I don't expect it. We are trying to put puzzle pieces together.*<br>2. *Some explanation, just an answer...toward why she was the way she was*<br>3. *Maybe some closure if there is an answer....* |
| Future Planning | 1. *One day down the road, we may have kids. Figure out what steps to take.*<br>2. *(I am) looking for help with future children, the chance of it recurring.*<br>3. *I want to know why our baby was so sick and if my husband or I carry a mutant gene that could cause this to happen with further children. Also if we are able to have more children in the future.* |
| Post-Sequencing |  |
| Uncertainty | 1. *This is a new field and it doesn't answer all the questions.*<br>2. *They weren't able to find anything specific but it could be something we still need to worry about.*<br>3. *I guess there's always a chance something could go wrong though.*<br>4. *We just don't really know what happened. We may never know.*<br>5. *There just isn't really an exact answer. I guess I'm in the same place as I was the last time we talked, I get it, and there isn't really an answer.* |
| Gratitude | 1. *I learned that learning the answer doesn't really change it. We are healed regardless of knowing the answer. I didn't even need it.*<br>2. *It gave us peace of mind, knowing that there wasn't anything obvious.*<br>3. *Thank you; there was nothing that we could have done to prevent what happened to our baby.*<br>4. *I feel like this was really important for me to be able to move on. The odds of happening again are probably slim. ...it closed a door on what happened. A relief, a gift.*<br>5. *We were grateful that no genetic issue was found in our baby.* |

## Discussion

Our study describes the unique experience of women utilizing trio-ES for prenatal genetic diagnosis. Short answer and survey responses obtained pre-sequencing demonstrate that the majority of women had high expectations of trio-ES to provide an answer related to their current pregnancy and also desired reassurance or answers about future pregnancies. Moreover, mean anxiety and depression scores within the cohort were significantly higher pre-sequencing than post; we hypothesize that this could be driven by the typical series of events required to obtain fetal genetic diagnosis, the burden of recent decision making regarding termination versus continuation of pregnancy, and, as suggested by the short answer responses, the uncertainty regarding future reproductive plans. It also may point to a role that information and contextualized support may have in mitigating anxiety. Importantly, the results speak to the significant mental health burden associated with bearing an anomalous pregnancy and the need for additional maternal support and mental health monitoring during the diagnostic process and after receipt of results.

Prior studies have demonstrated the substantial psychological repercussions for women in the setting of an anomalous pregnancy or pregnancy loss.^20 21^ Our results suggest that there may be additional mental health impacts from test-related anxiety and distress, particularly among women who receive results from testing. This may also be related to uncertainty of shared results and provision of secondary findings. Bernhardt et. al identified that uncertainty and unquantifiable .risks particularly impacted maternal psychological wellbeing after receiving microarray results for fetal genetic diagnosis. Ultimately, such results led to an anxiety-ridden pregnancy and postnatal period, increasing decisional conflict regarding testing and leading to results being viewed as “toxic knowledge.”^22^ While we noted differences in anxiety and depression scores related to study results, it is worth noting that scores decreased after completion of testing. This may relate to the fact that results were provided after decisions about pregnancy continuation were already made, which is in contrast to the study by Barnhardt et. al. Still, the results of our study in the context of prior literature suggest that uncertain results and secondary findings may impact long-term maternal psychological outcomes, particularly altering family dynamics, relationships, and support networks. Future studies need to target the longitudinal impact of next generation sequencing on parental psycho-social well-being in order to determine best practices for implementation and provision of adequate support.

Our study also demonstrates that nearly half of women in our cohort remained uncertain about future reproductive plans after trio-ES even though most women initially pursued advanced sequencing to assist with future reproductive planning. This highlights the importance of goals clarification in pretest counseling. Prior studies have suggested that given the increasingly common role of prenatal genetic screening and testing in the diagnostic algorithm, many women opt for the opportunity to receive additional information without carefully considering the risks and benefits of testing.^23 24^ Yet limiting pretest counseling to risks and benefits may not provide a true assessment of whether information provided will ultimately be psychologically beneficial or harmful. Instead, there is a significant need for goals clarification prior to pursuit of trio-ES and improvement in pre-test counseling to include uncertainty of results and how parents think they would fair in the face of unquantifiable risk.^22^

Post-sequencing, women who received any trio-ES result were more likely to experience test related anxiety and distress, test-related uncertainty, and had worse psychological adaptation to the genetic information provided compared to those that had no-result. Notably, in our study, there was no association between maternal demographic characteristics, baseline genetic knowledge and post-sequencing survey scores. As expected, baseline genetic knowledge was strongly associated with education level and previous experience with genetic testing. The lack of association between baseline genetic knowledge and post-sequencing survey results is in contrast to other literature that suggest heath literacy and medical knowledge are critically important to medical decision-making, reduction of decisional conflict, and adaptation of health information. ^14 25 26^ It is possible that the association was not easily identified given the homogeneity of our cohort, which was predominantly Non-Hispanic White, educated women. As such, future studies are necessary to determine how psychological outcomes may differ in populations that have lower health literacy or limited experience with genetic testing, or who approach genetic testing with different values or from different cultural contexts. It also brings forward the issue of representation in genetic studies and the importance of identifying barriers to a more diverse group of women electing to participate in the study.

The issues surrounding pre and post-test counseling, including interpretation of results and potential patient harms associated with anxiety, uncertainty, and missed parental expectations are in line with previously noted challenges of incorporating trio-ES into the prenatal fold.^27 28^ Previous work from our group discussed specific examples of ethical counseling challenges that have risen from this ongoing prospective cohort, identifying the importance of effective and appropriate communication of uncertainty and how it may impact long-term parental decisions.^7^ In particular, prior studies have identified the conflict between respect for patient autonomy and the potential harms of uncertainty and unquantifiable risk, ultimately resulting in outcomes that are discordant with robustly autonomous medical decision making.^29^ Prior studies have suggested that use of evidence-based counseling methods, guidelines on most applicable clinical scenarios for trio-ES, and standardization of reporting of results may improve patient autonomy in decision making ^27^

Our study has many strengths. It is a large, prospective cohort with longitudinal data. To our knowledge, we are the largest reporting study to date of maternal psychological outcomes after use of prenatal trio-ES. Our diagnostic yield is in line with prior studies. In our cohort, 22.3% of women received results from trio-ES that were suspected to explain fetal findings. Prior studies report a 10-57% diagnostic yield when using trio-ES^30 31^ Moreover, we used validated surveys and scales, adapted to assess test-related psychological outcomes in this particular scenario, and combined these efforts with qualitative methods for mixed methods results.

Yet, the findings of ours study should be interpreted within the context of its limitations. Our cohort is primarily composed of women that identify as Non-Hispanic White race, have college or greater education, and fall into high earning income groups. Thus, our results may not be generalizable to the general population. Future studies should work toward understanding and addressing barriers to enrolling women from diverse as well as marginalized populations in prenatal diagnosis studies and enroll a more representative population. These data will be critical in producing more generalizable results and inform a strategy for more equitable use of next-generation sequencing strategies. Women of lower socio-economic status, educational backgrounds, and from a wider range of ethnicities may have psychosocial outcomes with use of trio-ES that are significantly different from those captured in our study. Expanding our understanding of impact of trio-ES on diverse populations is integral to the development of an equitable and supportive platform to offer this technology.

Future studies should focus on the longitudinal psycho-social ramifications of advanced sequencing strategies, with the goal of developing resources to provide support during the pretest and post test periods. Examples include use of additional interactions with the prenatal genetic counselor via phone or web-based video platform or development of a pre-established psychological counseling support team for these women. The use of decision aids may also be a method of providing information while clarifying parental values and assessing ability to cope with uncertainty. Critically, such studies should focus on recruiting a diverse cohort in order to ensure that strategies for implementation are developed with a robust understanding the impact of advancing technology on parents of various ethnic, educational, and financial backgrounds. Advanced sequencing strategies offer significant prospect to improve the accuracy of prenatal genetic diagnosis. With continued growth in our understanding of parental psychological needs we will be able to improve the manner in which these technologies are offered and provide benefit to parents seeking answers.

## Data Availability

Data Sharing: The data that support the findings of this study are available on request from the corresponding author. The data are not publicly available due to privacy or ethical restrictions.

## Data Sharing

The data that support the findings of this study are available on request from the corresponding author. The data are not publicly available due to privacy or ethical restrictions.

